# ACTIVATE-2: A DOUBLE-BLIND RANDOMIZED TRIAL OF BCG VACCINATION AGAINST COVID19 IN INDIVIDUALS AT RISK

**DOI:** 10.1101/2021.05.20.21257520

**Authors:** Maria Tsilika, Esther Taks, Konstantinos Dolianitis, Antigone Kotsaki, Konstantinos Leventogiannis, Christina Damoulari, Maria Kostoula, Maria Paneta, Georgios Adamis, Ilias C. Papanikolaou, Kimon Stamatelopoulos, Amalia Bolanou, Konstantinos Katsaros, Christina Delavinia, Ioannis Perdios, Aggeliki Pandi, Konstantinos Tsiakos, Nektarios Proios, Emmanouela Kalogianni, Ioannis Delis, Efstathios Skliros, Karolina Akinosoglou, Aggeliki Perdikouli, Garyfallia Poulakou, Haralampos Milionis, Eva Athanassopoulou, Eleftheria Kalpaki, Leda Efstratiou, Varvara Perraki, Antonios Papadopoulos, Mihai G. Netea, Evangelos J. Giamarellos-Bourboulis

## Abstract

BCG vaccination induces heterologous protection against respiratory tract infections, and in children improves survival independently of tuberculosis prevention. The phase III ACTIVATE-2 study assessed whether BCG could also protect against COVID19 in the elderly. In this double-blind, randomized trial, elderly Greek patients were randomized (1:1) to receive either BCG revaccination or placebo at hospital discharge, followed by 6 months observation for incidence of COVID19 infection. BCG revaccination resulted in 68% risk reduction for total COVID19 clinical and microbiological diagnoses (OR 0.32, 95% CI 0.13-0.79). Five patients in the placebo group and one in the BCG-vaccinated group had severe COVID19 that necessitated hospitalization. 3 months after BCG vaccination 1.3% of placebo and 4.7% of BCG-vaccinated volunteers had anti-SARS-CoV-2 antibodies. These data argue that BCG revaccination is safe and protects the elderly against COVID19. BCG revaccination may represent a viable preventive measure against COVID19.

## INTRODUCTION

In December 2019, a novel enveloped RNA betacoronavirus was detected in a patient with pneumonia in Wuhan, the capital city of Hubei province of China (Huang et al., 2020). The virus that was subsequently isolated and sequenced was named the severe acute respiratory syndrome coronavirus-2 (SARS-CoV-2), and the disease it causes coronavirus disease-19 (COVID-19). SARS-CoV-2 spread around the world since the beginning of 2020, causing a pandemic that brought suffering and death to millions of people, and has caused the most severe healthcare emergency in the world since World War II.

The most important preventive tool against a new infection are vaccines, and a concerted effort has been launched around the world for the development of COVID-19 vaccines. As the beginning of 2021, a mere one year later, several new and effective vaccines against this new infection have been developed, based on both old (inactivated virus, recombinant proteins) and new (mRNA technology, adenovirus vectors) platform technologies (Forni and Mantovani, 2021). Vaccination programs with several of these vaccines have been initiated, and the beneficial impact of vaccination on the pandemic in countries that managed to vaccinate a significant proportion of the population is already starting to be felt (Amit et al., 2021). Unfortunately, the design and development of successful specific vaccines is time-consuming, with shortages of vaccines being encountered in the majority of countries: it is expected that 2 to 3 years will be needed until vaccines will be sufficiently available to all countries.

These inherent challenges posed by the development of completely new vaccines against a new pathogen argue for the identification of alternative preventive approaches that could at least partially protect the population until specific vaccines are developed and become available. One such approach that has been proposed for the prevention of COVID19 is the use of live-attenuated vaccines with known protective effects against heterologous infections (Netea et al., 2020). Bacillus Calmette-Guérin (BCG) was developed as a vaccine against tuberculosis, but epidemiological studies have suggested its capability to protect also against other infectious diseases (Benn et al., 2013). In infants, early administration of BCG vaccination leads to reduced child mortality, mainly as a result of reduced neonatal sepsis and respiratory infections (Aaby et al., 2011; Biering-Sørensen et al., 2012; Prentice et al., 2021), while revaccination with BCG in adolescents has shown a 70% decrease in the incidence of respiratory tract infections compared to placebo (Nemes et al., 2018). In addition, we also recently demonstrated in a randomized placebo-controlled trial (ACTIVATE), that BCG revaccination in elderly individuals can induce up to 80% reduction of the incidence of respiratory tract infections (Giamarellos-Bourboulis et al., 2020).

These earlier observations have led to the hypothesis that BCG vaccination may induce protection against COVID-19 as well (O’Neill and Netea, 2020). This hypothesis seems to be supported by a number of ecological studies that suggest an association between childhood vaccination and a low prevalence and severity of COVID-19 in various countries (Berg et al., 2020; Escobar et al., 2020). Retrospective studies of BCG vaccinated individuals have also shown that BCG is safe in the context of COVID-19 (Moorlag et al., 2020), and seems to be associated with protection against the infection or its severity (Rivas et al., 2021). Based on these intriguing arguments for a potential beneficial effect of BCG against COVID-19, a number of clinical prospective trials have been initiated around the world to investigate the effects of BCG vaccination (Junqueira-Kipnis et al., 2020; Madsen et al., 2020; Ten Doesschate et al., 2020). No data are yet available regarding the effect of BCG vaccination in the controlled settings of a randomized controlled trial.

The ACTIVATE-2 study is a placebo-controlled phase III trial for the investigation of the impact of BCG vaccination against COVID-19 in individuals at high-risk for COVID-19. High-risk was defined 50 years or more with comorbidities. In a cohort of individuals at high-risk from Greece, we observe that BCG revaccination results in an 68% reduction of the relative risk to develop COVID-19, compared with placebo-vaccinated controls, during a 6-months follow-up. In the placebo group five patients needed hospitalization due to COVID-19 complications, while only one patient in the BCG revaccination group had severe COVID19. These data suggest a potentially important role of BCG revaccination as a complementary tool against the pandemic.

## RESULTS

### Baseline characteristics are comparable between the two arms of vaccination

516 eligible participants were screened between June 2020 to October 2020. The first participant was enrolled on June 6, 2020 and the follow-up of the last participant was completed on April 19, 2021. No other people were willing to participate after October 2020 as the start of the regular vaccination programs for the general population has already been announced. Among them, 215 individuals were excluded due to positive skin tuberculin test (n=208), hematological malignancies (n=3), positive COVID19 serology, steroids, solid malignancy and chemotherapy (n=1 for each condition). 301 patients were randomized to double-blind vaccination: 153 individuals received placebo, and 148 individuals received BCG (Figure 1). 3-months interim analysis included 153 patients allocated to placebo vaccination and 148 patients allocated to BCG vaccination. Baseline characteristics were similar between the two arms (Table 1). 55 individuals were lost to follow-up in the placebo group, and 56 individuals in the BCG vaccination group after the first three months; as such, 6-month analysis was performed in 98 placebo-vaccinated individuals and 92 BCG-vaccinated individuals.

**Table 1.** Demographics of enrolled patients in the placebo and BCG vaccinated groups.

|  | Placebo (n=152) | BCG (n=148) | p-value |
| --- | --- | --- | --- |
| Male gender, n (%) | 106 (69.7) | 98 (66.2) | 0.538 |
| Age (years), mean (SD) | 68.7 (10.6) | 68.6 (10.4) | 0.912 |
| Charlson's comorbidity index | 3.86 (1.53) | 3.66 (1.52) | 0.112 |
| Comorbidities, n (%) |  |  |  |
| Coronary heart disease | 24 (15.8) | 17 (11.5) | 0.315 |
| Chronic heart failure | 10 (6.6) | 5 (3.4) | 0.290 |
| Type 2 diabetes mellitus | 33 (21.7) | 28 (18.9) | 0.569 |
| Vascular hypertension | 49 (32.2) | 38 (25.7) | 0.252 |
| Peripheral vascular disease | 19 (12.5) | 12 (8.1) | 0.256 |
| Chronic renal disease | 4 (2.6) | 1 (0.7) | 0.371 |
| Stroke | 9 (5.9) | 3 (2.0) | 0.138 |
| Chronic obstructive pulmonary disease | 35 (23.0) | 38 (25.7) | 0.687 |
| Hypothyroidism | 5 (3.3) | 6 (4.1) | 0.768 |
| Any surgery | 36 (23.7) | 39 (26.5) | 0.597 |
| Recent hospitalization | 20 (13.2) | 20 (13.5) | 1.00 |

**Figure 1.**
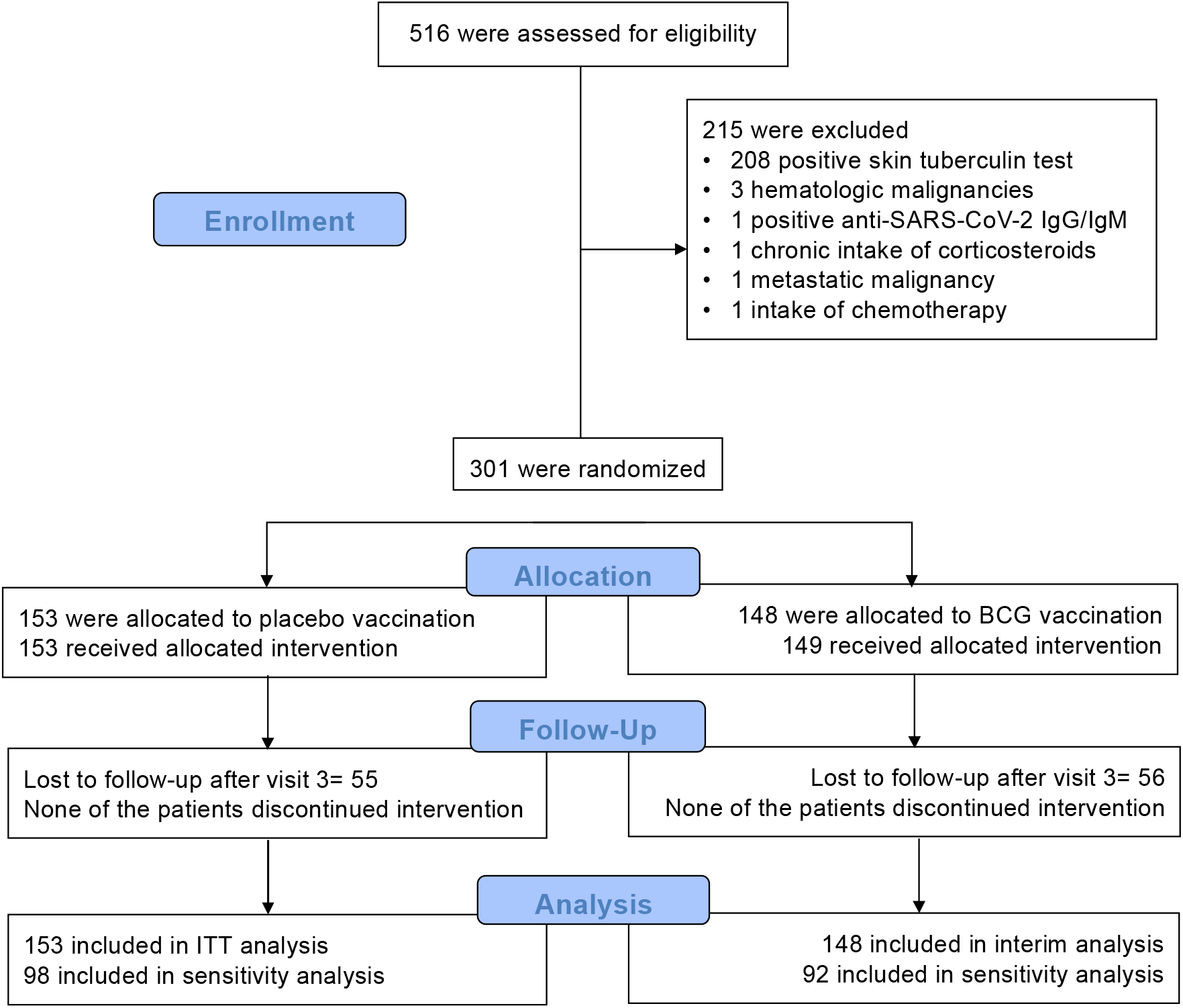
Study flow chart of ACTIVATE-2 study.

**Figure 2.**
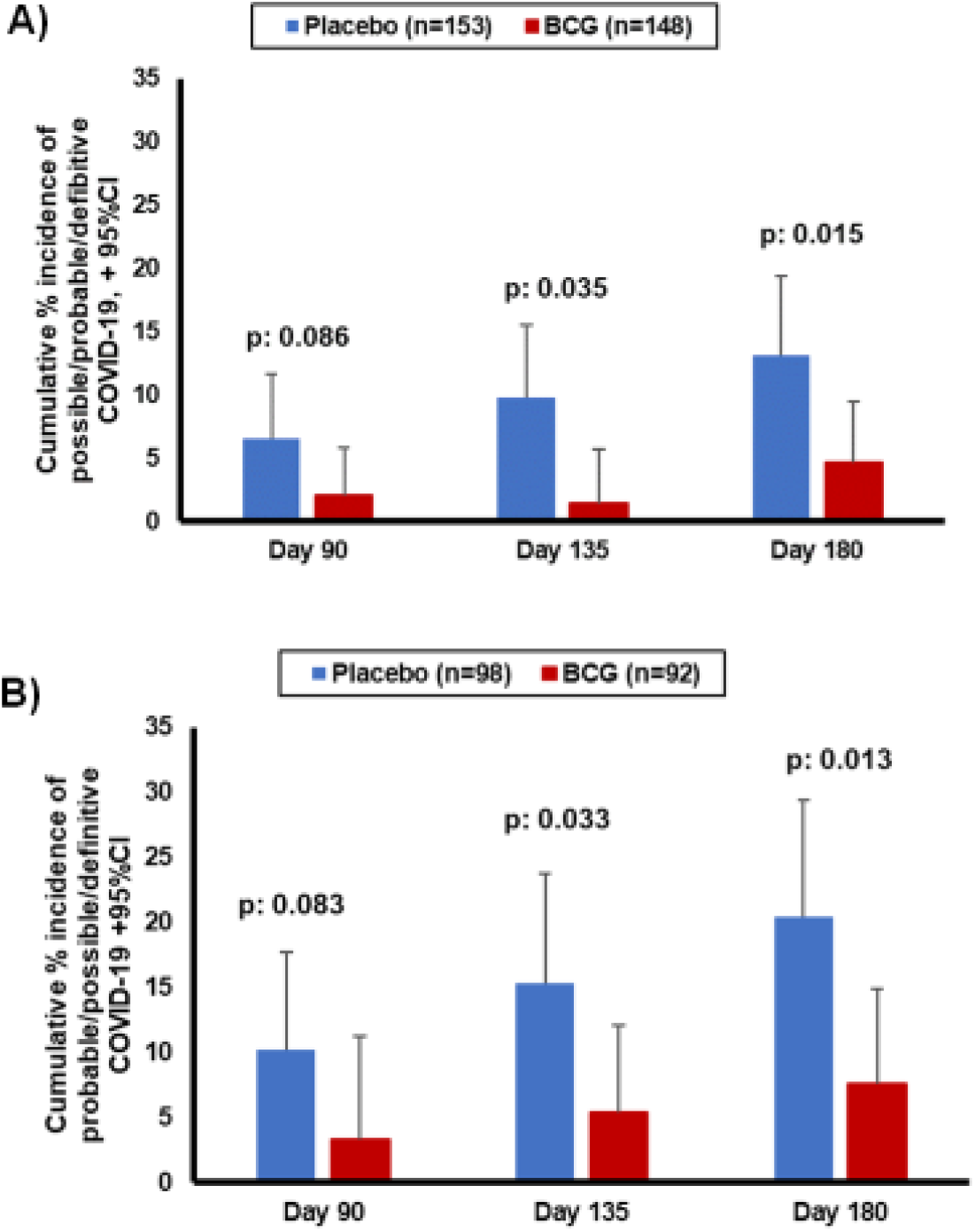
Cumulative incidence of probable/possible/definitive COVID-19 diagnosis during the study in the entire cohort (A) or in the patients that have completed the 180 days follow-up (B)

### BCG vaccination decreases the incidence of COVID-19

The primary outcome of the trial was incidence of a new COVID-19-possible/probable/definitive infection during the first 3-month period of follow-up after vaccination. This was a composite endpoint that included symptoms compatible with COVID-19 and/or the microbiological diagnosis of COVID-19. During these first 3 months after the vaccination the overall incidence of COVID-19 in Greece was low, and thus the number of COVID-19 diagnoses was low in both groups (10 patients in placebo vs. two patients in BCG group, p=0.086). In contrast, 6-months after vaccination, the total number of COVID-19 diagnoses (possible/probable/definitive) was significantly lower in the BCG-vaccinated group compared with the placebo group: OR 0.32 in multivariate analysis (95% CI 0.13-0.79, p=0.014) (Table 2).

**Table 2.** Univariate and multivariate analysis of variables associated with the overall incidence of COVID-19 compatible symptoms and/or definitive COVID-19.

|  | COVID-19 positive questionnaire and/or definitive COVID-19 |  | Univariate analysis |  | Multivariate analysis |  |
| --- | --- | --- | --- | --- | --- | --- |
|  | No (n=274) | Yes (n=27) | OR (95%CI) | p-value | OR (95%CI) | p-value |
| BCG vaccination, n (%) | 141 (51.5) | 7 (25.9) | 0.33 (0.14-0.81) | 0.015 | 0.32 (0.13-0.79) | 0.014 |
| Coronary heart disease, n (%) | 35 (12.8) | 7 (25.9) | 2.39 (0.94-6.06) | 0.067 | * |  |
| Type 2 diabetes mellitus, n (%) | 52 (19.0) | 10 (37.0) | 2.51 (1.09-5.80) | 0.031 | ** |  |
| Recent hospitalization, n (%) | 33 (12.0) | 8 (29.6) | 3.08 (1.25-7.58) | 0.015 | 3.16 (1.26-7.95) | 0.014 |

\*Variables not entering the equation because of no significance in the univariate analysis
\*\*Variables not entering the equation after two steps of forward step-wise analysis
Abbreviations CI: confidence intervals; OR: odds ratio

Definitive diagnosis of severe COVID-19 requiring hospitalization was present in 5 individuals in the placebo group and only one in the BCG group.

### Safety analysis

The number of treatment-emergent adverse events (TEAEs) was low and did not differ significantly between the placebo and BCG-vaccinated individuals, with the exception of erythema at the injection site which was more often reported in the BCG-vaccinated group, as expected (Table 3). None of the patients developed tuberculosis of systemic BCG-itis.

**Table 3.** Serious and non-serious treatment-emergent adverse events (TEAEs). The events that are related to COVID-19-related symptoms are not reported in this Table.

|  | Placebo (n=152) | BCG (n=148) | p-value |
| --- | --- | --- | --- |
| Total serious TEAEs, n (%) | 4 (2.6) | 1 (0.7) | 0.371 |
| Deaths | 3 (1.9) | 0 (0) | 0.247 |
| Urinary tract infection | 1 (0.7) | 0 (0) | 1.00 |
| Upper GI tract bleeding | 0 (0) | 1 (0.7) | 1.00 |
| Total non-serious TEAEs | 3 (2.0) | 10 (6.8) | 0.049 |
| Vaginal candidiasis | 1 (0.7) | 0 (0) | 1.00 |
| Hot flashes | 1 (0.7) | 0 (0) | 1.00 |
| Pain at the injection site | 0 (0) | 2 (1.4) | 0.242 |
| Erythema at the injection site | 0 (0) | 5 (3.4) | 0.028 |
| Pustule at the injection site | 1 (0.7) | 2 (1.4) | 1.00 |
| Hyperparathroidism | 0 (0) | 1 (0.7) | 1.00 |

### COVID-19 serology in placebo- and BCG-vaccinated individuals

At 3-months after vaccination, 300 volunteers donated blood for the assessment of specific antibodies against SARS-CoV-2, in order to diagnose asymptomatic infections. Interestingly, although the difference observed between the groups did not reach statistical significance, positive COVID-19 serology was measured in only 1.3% (2/153) individuals who received placebo, compared to 4.7% (7/148) volunteers who received BCG vaccine (p=0.099).

## DISCUSSION

In this study we present the data of the first phase III placebo-controlled randomized trial of BCG vaccination against COVID-19 to be completed. BCG vaccination resulted in a significantly reduced incidence of possible/probable/definitive COVID-19 in an elderly population 6-months after vaccination, compared to the placebo group. Although the number of severe infections was small, hospitalization due to severe COVID-19 occurred in five individuals vaccinated with placebo, but in only in one of the BCG-vaccinated elderly volunteers. These data argue for a potentially important role of BCG revaccination in the fight against the COVID-19 pandemic.

The ACTIVATE-2 trial was initiated based on the capacity of BCG to: i. reduce the incidence of respiratory tract infections in children and adults (Aaby et al., 2011; Biering-Sørensen et al., 2012; Giamarellos-Bourboulis et al., 2020; Nemes et al., 2018; Prentice et al., 2021); ii. exert antiviral effects in experimental models (Spencer et al., 1977); iii. reduce viremia in an experimental human model of viral infection (Arts et al., 2018); and the demonstrated safety in the context of the COVID-19 pandemic (Moorlag et al., 2020). We hypothesized that BCG vaccination may induce (partial) protection against susceptibility to and/or severity of SARS-CoV-2 infection, and this hypothesis was confirmed by the clinical data showing 68% reduction of COVID-19 retrospective diagnoses in the vaccinated individuals. While the data of the current study are in line with a number of studies showing an association between countries that implement BCG vaccination at birth and a low prevalence of COVID19 (Berg et al., 2020; Escobar et al., 2020), not all studies were however able to reproduce these findings (Hamiel et al., 2020; Wassenaar et al., 2020). Ecological studies are also prone to bias, and thus such arguments based on ecological analyses need to be considered with caution. On the other hand, other recent studies have also shown reduced COVID-19 incidence in an observational study of recent BCG vaccination (Amirlak et al., 2020), and a reduced severity of the disease in children vaccinated with BCG (Chen et al., 2021), all which support the observation reported here of a protective effect of BCG.

It has been recently demonstrated that the non-specific beneficial effects of BCG vaccination are due to epigenetic and metabolic reprogramming of innate immune cells such as myeloid cells and NK cells, leading to an increased antimicrobial activity, a process termed ‘trained immunity’ (Netea et al., 2016). Upon stimulation with certain microorganisms, the innate immune system becomes primed and is able to react faster and more efficient to secondary (and non-related) stimuli. In experimental studies, BCG has been shown to protect not only against bacterial and fungal infections, but against viral infections such as influenza as well (Spencer et al., 1977). Furthermore, we recently showed that in volunteers receiving yellow fever vaccine virus, those who had recently received BCG had – compared to placebo-treated subjects – less viremia, and improved anti-viral responses (Arts et al., 2018). The reduction in viremia correlated with myeloid-dependent trained immunity responses, rather than specific lymphocyte-dependent immune memory (Arts et al., 2018). The potential important role of trained immunity for the protection against COVID-19 is supported by the observation of enhanced innate immune responses post-vaccination in our recent ACTIVATE trial as well (Giamarellos-Bourboulis et al., 2020).

An intriguing observation was made when COVID-19 serology was investigated, in order to identify the potential asymptomatic cases: only 1.3% of the placebo-vaccinated individuals were positive, while 4.7% of the BCG-vaccinated individuals showed positive anti-SARS-CoV-2 antibodies. While the difference was not statistically significant due to the low numbers and it may be just by accident, it is tempting to speculate that this is a real observation: earlier studies have demonstrated that BCG improves serological responses to other vaccines (Leentjens et al., 2015; Ritz et al., 2013), and it may induce similar effects in asymptomatic COVID-19 infections. It has been demonstrated that mild or asymptomatic COVID-19 infections induce low antibody responses (Bölke et al., 2020; Kutsuna et al., 2020; Terpos et al., 2020; Yang and Ibarrondo, 2020): in contrast, a previous BCG vaccination may improve this and indirectly induce in an increased protection against the infection in individuals asymptomatically infected with SARS-CoV-2. Future studies are warranted to explore and validate this hypothesis.

Studies that investigated BCG vaccination have been recently initiated in a number of countries around the world both in children and adults. It is interesting to observe that a certain pattern starts to emerge in which trials in developing countries or countries that employed BCG vaccination programs show beneficial effects of BCG re-vaccination, while countries in which first BCG vaccination is administered in naïve populations of developed countries the effect seems to be less strong. In children, clinical trials in Africa have shown significant protection of BCG vaccination against mortality (Aaby et al., 2011; Biering-Sørensen et al., 2012) and infections (Prentice et al., 2021), but such effect failed to materialize in a recent large trial in Denmark (Stensballe et al., 2019). Interestingly however, in the Calmette study from Denmark, BCG vaccination induced a 30% protection of the child against infections if the mother was also BCG-vaccinated, while no effect was seen if the mother never received BCG (Berendsen et al., 2020). Similarly, BCG revaccination in South-African adolescents resulted in 73% less infections (Nemes et al., 2018), while BCG vaccination in Greek elderly resulted in 80% less respiratory tract infections (Giamarellos-Bourboulis et al., 2020). While we do not have information regarding the BCG vaccination status of the volunteers in the ACTIVATE-2 trial, Greece had a policy of BCG vaccination at birth in the temporal interval when the volunteers from this trial were born. The positive data in the present study contrasts the results of a recent interim analysis of the BCG-PRIME study from the Netherlands that suggested lack of significant effects of BCG vaccination (UMC Utrecht, 2021). Altogether, these data suggest important differences in the protective effects of BCG revaccination vs. BCG first vaccination, and these need to be investigated in future studies.

The present study also has limitations. The most important limitation is the small size of the study. Although the strong impact of BCG vaccination allowed to reach important conclusions regarding the effect on COVID-19, larger studies that are currently under way in different countries need to confirm the data of ACTIVATE-2. Another limitation is represented by the lack of microbiological testing in all patients with a clinical diagnosis of possible or probable COVID-19. Although we cannot exclude an alternative diagnosis is some of these patients, the context of the pandemic during tight societal restrictions makes COVID-19 diagnosis the most likely. Because of the small size of the study, no conclusions can be drawn regarding the effect of BCG vaccination on the severe forms of COVID-19, although the difference of hospitalizations between placebo and BCG groups suggests potential protection of the vaccination. A fourth limitation of the study is the lack of information regarding the SARS-CoV-2 strains that caused the infections in this study, and future studies need to investigate this aspect as well. Finally, we have limited information regarding the impact of BCG on the specific immunological host defense pathways against SARS-CoV-2 (specific antibodies and T-cell responses), and additional investigations should investigate these effects in more detail.

When considering the availability of the novel specific anti-SARS-CoV-2 vaccines, one could ask why do we need trials investigating the effect of BCG against COVID-19. There are several important reasons why such studies are still very important. First, the availability of the specific vaccines is limited, especially in developing countries: as the majority of developing countries have programs of neonatal BCG vaccination, a strategy using BCG revaccination could have important impact on the COVID-19 pandemic until the specific vaccines are available. Second, our ability to control the COVID-19 pandemic is continuously challenged by the appearance of new variants of the virus, some of them that show increased escape rates against the newly developed vaccines (Kustin et al., 2021). The employment of trained immunity-based vaccines that boost anti-viral host defense in an antigen-independent manner could thus prove very important in situations in which such viral variants become most prevailing. Third, the principle that heterologous vaccines can be employed with success in the real-life situation of a pandemic is extremely important for future pandemic preparedness: it is conceivable that a strategy employing such vaccines can be used as ‘bridge vaccination’ for partial protection of the population even before specific vaccines are available (Netea et al., 2020). It is to be hoped that the results of the present study will encourage more research for identifying the most effective set of heterologous vaccines that can be stored, tested, and used in a future pandemic to avoid the high healthcare, societal and economic toll that can be exerted by a dangerous new pathogen.

## Data Availability

The data are available upon reasonable request

## Acknowledgements

The study was funded in part by the Hellenic Institute for the Study of Sepsis and in part by Fast Grants of Emergent Ventures at the Mercatus Center, George Mason University.

## Author contributions

MT collected clinical data and maintained the study database, drafted the manuscript and gave final approval of the final version to be submitted.

ET contributed to data analysis. drafted the manuscript and gave final approval of the final version to be submitted.

KD, MK, GA, IP, KS, KK, CD, IP, KT, NP, EK, ID, ES, KA, AP, GP, HM and EA collected clinical data, reviewed the manuscript and gave final approval of the final version to be submitted.

MGN conceptualized the study, analysed the data, drafted the manuscript and gave approval of the final version to be submitted

EJGB designed the study protocol, analysed the data, drafted the manuscript and gave approval of the final version to be submitted

## STAR Methods

### Lead Contact

Information and requests for resources should be directed to and will be fulfilled by the Lead Contact, Evangelos J. Giamarellos-Bourboulis.

### Materials Availability

This study did not generate new unique reagents.

### Data and Code Availability

Data of this study are available after communication with the Lead Contact. A material transfer agreement will be needed.

## EXPERIMENTAL MODEL AND SUBJECT DETAILS

### ACTIVATE-2 clinical trial

ACTIVATE-2 (<u>A</u> randomized <u>c</u>linical trial for enhanced trained immune responses through Bacillus Calmette-Guérin <u>va</u>ccination to prevent inf<u>e</u>ctions by COVID-19) is a prospective, double-blind, randomized and placebo-controlled phase III clinical trial conducted among the general population in 11 departments of Internal Medicine in Greece. The protocol and its subsequent amendments were approved by the National Ethics Committee (approval 52/20) and by the National Organization for Medicine of Greece (approval IS 045-20) (EudraCT number 2020-002448-21; Clinicaltrial.gov NCT04414267). The trial was sponsored by the Hellenic Institute for the Study of Sepsis. The funders have no role in the design, conduct, analysis and interpretation of data, and decision to publish. The database lock was done on April 28th 2021. The investigators remained blind to the intervention until the results of the analysis became known.

Study participants were informed about the study by either announcement in the media or through advertisements that were put on each of the 11 participating hospitals. Enrolled participants should be of either gender, aged 50 years or older and have history of coronary heart disease of or chronic obstructive pulmonary disease or have Charlson’s comorbidity index (CCI) more than 3. Enrolled people should also have skin tuberculin test diameter less than 10mm and negative serum testing for immunoglobulin G and M against SARS-CoV-2. Exclusion criteria were: infection by the Human Immunodeficiency Virus-1 (HIV-1); primary immunodeficiency; solid organ transplantation; bone marrow transplantation; intake of chemotherapy the last two months; intake of radiotherapy the last two months; active hemalogical or solid tumor malignancy; intake of any anti-cytokine therapies; intake of oral or intravenous steroids defined as daily doses of 10mg prednisone or equivalent for longer than the last 3 months. All participants provided written informed consent before enrolment.

Assessment of eligibility was done after thorough study of the patient past history. For eligible participants, skin tuberculin test was done by intradermal injection of 0.1ml of tuberculin in one forearm. Participants were asked to return after 48 to 72 days and in the case of diameter of the test less than 10mm they were subject to measurement of anti-SARS IgG/IgM antibodies. Antibodies were measured using the WIZ Biotech rapid measurement kit and employing single drops of blood from the forefingers. Participants negative for antibodies were allowed to be vaccinated. Vaccination was done with the subdermal injection of 0.1ml of sodium chloride 0.9% or with 0.1ml of BCG vaccine. The studied BCG Vaccine is a live freeze-dried vaccine derived from attenuated strain of *Mycobacterium bovis* (Bacillus Calmette Guerin Moscow strain 361-I) and each 0.1 ml contains 2 to 8× 10^5^ colony forming unis. The intradermal injection of NaCl (placebo) or BCG (active vaccine) was done in the deltoid region.

Following vaccination patients were asked to return at the study site for four more times (study visit 2 on days 45 ±5 from date of visit 1; study visit 3 on days 90 ±5 from date of visit 1; study visit 4 on days 135 ±5 from date of visit 1; and study visit 5 on days 180 ±5 from date of visit 1). Study visits 2, 5 and 6 could also be phone visits. On each visit patients were thorough asked for any adverse events (AE). Then they were asked if they had suffered from COVID-19 diagnosed with molecular test. These cases were considered as patients with definitive COVID-19. Then participants were asked to provide answers to a questionnaire if they had experienced COVID-19-related symptoms. The combinations of answers allowed to classify them as sufferers from possible or probable COVID-19. Participants were also subject to a second rapid test for anti-SARS IgG/IgM antibodies on visit 3.

The primary outcome was assessed on visit 3 (90 ± 5 days from the date of visit 1) as the incidence of total cases of possible/probable/definitive COVID-19 the first 90 ± 5 days post vaccination. The incidence of total cases of possible/probable/definitive COVID-19 the first 135 ± 5 days post vaccination and the first 180 ± 5 days post vaccination were the secondary study endpoints (visits 4 and 5 respectively).

TEAEs and Serious TEAEs were captured from baseline until the last patient’s evaluation. An adverse event was defined as any undesirable medical occurrence in a subject administered a pharmaceutical product and which does not necessarily have a causal relationship with this treatment. The adverse event could be a sign, a symptom, or an abnormal laboratory finding. AEs meeting any of the following criteria were considered SAEs: death; life-threatening situation; inpatient hospitalization or prolongation of existing hospitalization; persistent or significant disability/incapacity; congenital anom- aly/birth defects; important medical events/experiences; and spontaneous and elective abortions experienced by study subject. All others AEs were reported as non-serious. All AEs were graded as: mild (transient and well-tolerated by the patient), moderate (causing discomfort and affecting the usual activities of the patient) and severe (affecting the usual activities to an important de- gree and causing disability or are life-threatening). The relationship of the AE to the study drug was reported as probably-related, possibly-related; probably not-related and unrelated

## Statistical analysis

Results of qualitative variables were expressed as percentages and 95% confidence intervals (CIs) and of quantitative variables as means and standard error. Comparisons between placebo-vaccinated and BCG vaccinated individuals was done by the Fisher’s exact test and by the Student’s t-test respectively. Step-wise logistic regression analysis was done to investigate the variables associated with probable/possible/definitive COVID-19 until visit 5. The COVID-19 cases entered the equation as dependent variables and variables defined by the univariate analysis to be associated with COVID-19 as independent variables. The odds ratio (OR) and the 95% CIs were defined.

